# Ownership and COVID-19 in care homes for older people: A living systematic review of outbreaks, infections, and mortalities

**DOI:** 10.1101/2021.01.28.21250547

**Authors:** Anders Malthe Bach-Mortensen, Ben Verboom, Ani Movsisyan, Michelle Degli Esposti

## Abstract

**Background:** The adult social care sector is increasingly outsourced to for-profit providers, who constitute the largest provider of care homes in many developed countries. During the COVID-19 pandemic, for-profit providers have been accused of failing their residents by prioritising profits over care, prevention, and caution, which has been reported to result in a higher prevalence of COVID-19 infections and deaths in for-profit care homes. Although many of these reports are anecdotal or based on news reports, there is a growing body of academic research investigating ownership variation across COVID-19 outcomes, which has not been systematically appraised and synthesised.

**Objectives:** To identify, appraise, and synthesise the available research on ownership variation in COVID-19 outcomes (outbreaks, infections, deaths, shortage of personal protective equipment (PPE) and staff) across for-profit, public, and non-profit care homes for older people, and to update our findings as new research becomes available.

**Design:** Living systematic review.

**Methods:** This review was prospectively registered with Prospero (CRD42020218673). We searched 17 databases and performed forward and backward citation tracking of all included studies. Search results were screened and reviewed in duplicate. Risk of bias (RoB) was assessed in duplicate according to the COSMOS-E guidance. Data was extracted by ABM and independently validated. The results were synthesised by country, RoB, and model adjustments, and visualised using harvest plots.

**Results:** Twenty-nine studies across five countries were included, with 75% of included studies conducted in the Unites States. For-profit ownership was not consistently associated with a higher probability of a COVID-19 outbreak. However, there was compelling evidence of worse COVID-19 outcomes following an outbreak, with for-profit care homes having higher rates of accumulative infections and deaths. For-profit providers were also associated with shortages in PPE, which may have contributed to the higher incidence of infections and deaths in the early stages of the pandemic. Chain affiliation was often correlated with an increased risk of outbreak but was usually not reported to be associated with higher rates of deaths and infections.

**Conclusion:** For-profit ownership was a consistent risk factor for higher cumulative COVID-19 infections and deaths in the first wave of the pandemic. Thus, ownership and the characteristics associated with FP care home providers may present key regulatable factors that can be addressed to improve health outcomes in vulnerable populations and reduce health disparities. This review will be updated as new research becomes published, which may change the conclusion of our synthesis.

## Introduction

The COVID-19 pandemic has disproportionately affected people living in residential care, who are estimated to account for approximately half of all COVID-19 deaths in developed countries ^1–3^. This disproportionate impact can be understood, in part, in terms of the vulnerability of people residing in care homes and a lack of early intervention and support ^4^. However, the structural and institutional risk factors are not yet well understood. In many countries, adult social care services are delivered by a combination of for-profit, non-profit, and public providers ^5^. It is well documented that the outsourcing of social care has significantly increased the market share of private, and in particular, for-profit care providers ^6,7^, which has motivated a large body of research investigating the association between care home ownership and quality of care ^8^. For example, a 2009 systematic review of 82 studies on ownership variation among nursing homes found that non-profit providers typically delivered higher quality services compared to for-profit providers^9^; a finding that has since been replicated ^6,7,10^.

During the COVID-19 pandemic, some news coverage expressed concern that for-profit providers have failed their residents by prioritising profits over care, prevention, and caution ^11^, resulting in reports of higher rates of COVID-19 infections and deaths in for-profit care homes ^12,13^. Although many of these reports are not peer-reviewed – located, for example, in newspaper publications – there is a growing body of academic research that investigates ownership variation across COVID-19 outcomes, such as outbreaks, infection rates, and mortality ^14,15^.

The COVID-19 pandemic has tested the capability of not only individual care homes, but also the commissioning system in which they operate. It is known that funding for adult social care has been characterised by years of austerity ^16^ at the expense of staffing, quality, and support, which may have been detrimental to the ability of care homes to cope with the pandemic ^17^. As such, the experiences of the adult social care sector during the pandemic must not be forgotten, and the growing body of research on ownership variation across COVID-19 outcomes offers an important opportunity to revisit the impact of outsourcing social care services to for-profit providers.

### Research Question/objectives

The aim of this living systematic review is to identify, appraise, and synthesise the available research on ownership variation in outbreaks and infections across for-profit, public, and non-profit care homes for older people, and to update our findings as new research becomes available. A review protocol was registered prospectively on OSF ^18^ and on Prospero (CRD42020218673).

## Methods

COVID-19 related research is published at a high frequency, and the time between submission and publication is substantially shorter than for non-COVID-19 research^19^, which makes it particularly important that this rapidly growing body of evidence is critically appraised and systematically synthesised regularly. To ensure that this review will represent a *recent* and *relevant* synthesis of the available evidence on the topic, it will be conducted as a living systematic review, which is defined as “*a systematic review that is continually updated, incorporating relevant new evidence as it becomes available*” ^20,21^. Specifically, we plan to update our results every 6 months for two years after initial publication. An update will entail (1) running the below search string for the updated time period, and (2) forward and backward citation tracking of all included studies. Any studies added throughout this process will be incorporated in the review and the synthesis, results, and interpretation will be adapted accordingly. This review is conducted and reported in accordance with PRISMA guidelines ^22^(see supplementary material for PRISMA checklist)

### Search strategy

Before developing our search strategy, we hand-searched Google Scholar and preprint repositories for relevant articles and performed citation searches on all identified articles. This preliminary sample of includable studies were used to design our search strategy and to test its specificity. The search strategy was piloted and adjusted until all pre-identified studies were identified. The full search strategy can be found in the supplementary material. Our search string was implemented in the following databases: ABI/INFORM Global, Coronavirus Research Database, Criminology Collection International (criminal justice database and NCJRS abstracts database), International Bibliography of the Social Sciences (IBSS), Politics Collection (PAIS index, policy file index, political science database, and worldwide political science abstracts), Social Science Database, Sociology Collection (applied social science index and abstracts, sociological abstracts, and sociology database) via ProQuest. We searched Embase, Global Health, Medline, and PsycINFO via Ovid. Two authors (ABM and MDE) double-screened all search results using Rayyan and subsequently reviewed the full text versions of potentially eligible studies. To ensure that the results of this review were as up-to-date as possible, we tracked all new research citing the studies included from our data base search every 2 weeks, with the latest one conducted on January 26^th^, 2021. These on-going forward citation searches led to the identification of 9 additional studies, most of which were published after our formal search was conducted (see PRISMA diagram for details).

### Inclusion criteria

We assessed study eligibility based on four criteria. *First*, studies had to investigate variation in COVID-19 outbreaks, infection rates, and/or excess or COVID-19 related mortalities or outcomes related to personal protective equipment (PPE) use and availability, staff shortage, preparedness, and infection and mortality among staff and visitors. We did not exclude studies based on how the COVID-19 outcomes were operationalised (e.g. if the infections were confirmed by PCR test or by self-report, and whether the analysed outcomes were dichotomised or continuous), although these aspects were considered in our risk of bias assessments. *Second*, studies had to investigate variation in any of the above outcomes across ownership categories, which is conventionally operationalised as ‘for-profit’ (e.g. private care homes run for-profit), ‘non-profit’ (e.g. registered private not-for-profit care home or charities), and ‘public’ (e.g. municipal or local authority care home). However, ‘ownership’ is not consistently operationalised in the literature, and terms such as ‘for-profit’, ‘non-profit’, ‘private’, ‘chain affiliated’, and ‘public’ are rarely clearly defined. Since the objective of this review is to appraise and synthesise research on ownership variation, we considered any definition or classification of ownership. However, the nuances of different ownership categorisations were considered in our synthesis and interpretation (see Table A1 in the appendix for a breakdown and description of different ownership categories). *Third*, studies had to employ an observational research design, including, but not limited to, cross-sectional studies, cohort studies, and secondary analyses of registry data. Both published and unpublished manuscripts (e.g., preprints and reports) were eligible for inclusion. *Fourth*, studies had to investigate residential care homes for older people, including, but not limited to, long-term care facilities, nursing homes, and retirement homes.

### Data extraction, (selection and coding)

Descriptive information on the citation details (author, title, journal) and study characteristics (research design, analysis, sample details) were retrieved from all included articles. We also extracted detailed information on the descriptive data (e.g., country, source, and period of data coverage) and the outcome and exposure variables (e.g., definition, operationalisation, and cut-offs). All results relating to ownership variation across COVID-19 related outcomes and accompanying interpretations were extracted for all studies. The results were extracted by ABM and independently validated by the co-authors.

### Risk of bias assessment

Risk of bias (RoB) was assessed using the COSMOS-E guidance ^23^. We employed this guidance rather than, for example, ROBINS-I for non-randomised intervention studies, as it is specifically designed for systematically reviewing observational and correlational research. Specifically, we evaluated the following bias domains: confounding, selection bias, and information bias. In line with the consistent recommendation to avoid quantitative scoring of risk domains^23,24^, all RoB assessments were based on the qualitative subjective assessment of the reviewers, which were decided through discussion and in consensus. All assessments were conducted with the focus of our review (variation in COVID-19 outcomes across ownership) in mind, and the assessments may thus not represent the risk of bias across other investigated associations and outcomes. The overall RoB assessment for each study was based on the lowest assessment in any bias domain. We did not exclude studies based on the RoB assessments.

### Data synthesis

Due to a high degree of heterogeneity among the included studies in terms of model specifications, operationalisation of outcomes, inconsistent ownership categorisation of the reference group, and overlapping data, we did not perform a statistical meta-analysis of the included results. This decision was made with attention to the pitfalls of employing statistical methods and assumptions designed for the analysis of very homogenous data or randomised controlled trials of interventions to observational and correlational research ^25^. If appropriate however, we will meta-analyse the results in later versions of this living review. Our synthesis was guided by the Synthesis Without Meta-analysis (SWiM) guidelines ^26^ and can be described as follows.

*First*, we narratively summarised key characteristics of the included studies, such as publication type, sample details, ownership categorisation, and data sources. *Second*, we assessed the risk of bias across all included studies. *Third*, we developed an overview of the strength of evidence for specific outcomes and model adjustments of all contributing studies. The model adjustment categories were developed according to aspects known to influence the relationship between ownership and care homes. *Fourth*, we constructed a harvest plot to graphically illustrate the direction of the effects across different outcomes and ownership categories with attention to the model specifications and risk of bias of contributing studies ^27^. Harvest plots serve as a way to synthesise and describe a heterogenous body of evidence, which cannot be meaningfully synthesised by means of a meta-analysis. *Last*, we analysed and examined the role or mediating risk associated to both FP ownership and COVID-19.

## Results

### Search results

Our search strategy yielded 3,845 results, of which 3,514 were left after removal of duplicates. These were double-screened by ABM and MDE, after which 81 were independently assessed in full-text. Of these, 20 studies were deemed eligible for inclusion. A forward citation search was conducted on this sample every two weeks until January 26^th^, 2021, in order to identify emerging new research on the topic. An additional 9 eligible studies (primarily preprints and government reports) were identified through this process. Twenty-nine studies were thus included in the first version of this review.

### Description of studies

The descriptive characteristics of all included research are shown in Table 1. Most of the included studies were peer-reviewed publications (24/29), with 2 government reports ^28,29^ and 3 preprints or working papers. The unit of analysis across all included studies was care homes. For-profit care homes were the largest ownership group across all studies that provided detailed sample information. All except for two studies ^30,31^ were published in July 2020 and after. Most studies were conducted in the United States (22/29), followed by Canada (3/29), England (2/29), Scotland (1/29), and France (1/29). Most included studies were considered cross-sectional and only three studies included more than one time-point in their analysis ^32–34^.

**Table 1:** Study and outcome characteristics

| Study | Type of study | Publication date | Research design | Country | Operationalisation of ownership | Includes chain? (y/n) | Sample size (care homes) | Sample size across ownership groups | COVID- 19 outcomes |
| --- | --- | --- | --- | --- | --- | --- | --- | --- | --- |
| (Bui et al., 2020) <sup>28</sup> | Government report | September 2020 | Cross-sectional | United States (West Virginia) | For-profit and non-profit | N | 123 | For-profit: 95 (77%)<br>Non-profit: 28 (23%) | Outbreaks (two or more cases) |
| (Chatterjee, Kelly, Qi, & Werner, 2020) <sup>43</sup> | Peer-reviewed publication | July 2020 | Cross-sectional | United States (23 States) | For-profit and non-profit | N | 8,943 | For-profit: 6473 (72%)<br>Non-profit: 2470 (28%) | Outbreaks (any cases) |
| (Abrams, Loomer, Gandhi, & Grabowski, 2020) <sup>44</sup> | Peer-reviewed publication | June 2020 | Cross-sectional | United states (30 States) | For-profit, non-profit and public | Y | 9,395 | For-profit: 6878 (73%)<br>Non-profit: 2178 (23%)<br>Government: 339 (4%) | Outbreaks (any cases) and number of infections |
| (Braun et al., 2020) <sup>35</sup> | Peer-reviewed publication | October 2020 | Cross-sectional | United States (unclear number of states) | Private-equity, for-profit non-profit, and public | Y | 11,470 | For-profit: 7793 (67.9%)<br>Non-profit: 2523 (22.0%)<br>Government: 511 (5.3%)<br>Private equity: 543 (4.7%) | Number of infections and deaths pr 1,000 residents, supply of PPE, and staffing shortage |
| (Yue Li, Temkin-Greener, Gao, & Cai, 2020) <sup>14</sup> | Peer-reviewed publication | June 2020 | Cross-sectional | United States (Connecticut) | For-profit and non-profit (including public providers) | Y | 215 | For-profit 179 (83%)<br>Non-Profit: 36 (17%) | Number of infections and deaths |
| (Yumeng Li, Fang, & He, 2020) <sup>45</sup> | Peer-reviewed publication | October 2020 | Cross-sectional | United States (unclear number of states) | For-profit, non-profit and public | Y | 14,062 | For-profit: 9,787 (70%)<br>Government: 903 (6.5%)<br>Non-profit: 3286 (23.5%) | Supply of PPE |
| (Stall, Jones, Brown, Rochon, & Costa, 2020) <sup>46</sup> | Peer-reviewed publication | August 2020 | Cross-sectional | Canada Ontario | For-profit, non-profit, and public | Y | 623 | For-profit: 360 (57.7%)<br>Non-profit: 162 (26.0%)<br>Government: 101 (16.2%) | Outbreaks (any cases), number of infections, and deaths |
| (He, Li, & Fang, 2020) <sup>47</sup> | Peer-reviewed publication | June 2020 | Cross-sectional | United States (California) | For-profit, non-profit, and public | N | 1,144 | For-profit: 956 (84%)<br>Government: 35 (3%)<br>Non-profit: 153 (13%) | Number of infections and deaths |
| (Sugg et al., 2021) <sup>48</sup> | Peer-reviewed publication | August 2020 | Cross-sectional | United States (unclear number of states) | For-profit, non-profit, and public | N | 13,709 | Unclear | Number of infections |
| (Dean, Venkataraman i, & Kimmel, 2020) <sup>49</sup> | Peer-reviewed publication | November 2020 | Cross-sectional | United States (NY state) | For-profit and non-profit | Y | 621 | Unclear | Deaths |
| (Harrington et al., 2020) <sup>50</sup> | Peer-reviewed publication | July 2020 | Cross-sectional | United States (California) | For-profit, non-profit (including public) | N | 1,091 | For-profit: 920 (84%)<br>non-profit & government: 171 (16%) | Outbreaks (any cases) |
| (Unruh, Yun, Zhang, Braun, & Jung, 2020) <sup>30</sup> | Peer-reviewed publication | June 2020 | Cross-sectional | United States (Connecticut, New Jersey, New York) | For-profit and non-profit | Y | 1,162 | Unclear | Deaths (dichotomised as 6 or more) |
| (Bowblis & Applebaum, 2020) <sup>32</sup> | Peer-reviewed publication | October 2020 | Cohort | United States (Ohio) | For-profit, non-profit, and public | Y | 942 | For-profit: 749 (79.4%)<br>Non-profit: 178 (19%)<br>Government: 15 (1.6%) | Outbreaks and number of infections (dichotomised as more than 20% of residents) |
| (Yue Li, Cen, Cai, & Temkin-Greener, 2020) <sup>39</sup> | Peer-reviewed publication | September 2020 | Cross-sectional | United States (unclear number of states) | For-profit, non-profit, and public | Y | 12,576 | For-profit: 8,861 (70%)<br>Non-profit: 2,938 (23%)<br>Government: 777 (7%) | Number of infections and deaths |
| (Fisman, Bogoch, Lapointe-Shaw, McCready, & Tuite, 2020) <sup>51</sup> | Peer-reviewed publication | July 2020 | Cross-sectional | Canada (Ontario) | For-profit, non-profit, municipal, and charitable | N | 627 | For-profit: 361 (57%)<br>Non-profit: 117 (18%)<br>Municipal: 101 (16%)<br>Charitable: 57 (9%) | Outbreaks (any cases) |
| (Xu, Intrator, & Bowblis, 2020) <sup>37</sup> | Peer-reviewed publication | August 2020 | Cross-sectional | United States (unclear number of states) | For-profit, non-profit, and public | Y | 11,920 | For-profit: 8561 (72%)<br>government: 647 (5%)<br>Not-for-profit: 2712 (23%) | Shortage of staff |
| (Gorges & Konetzka, 2020) <sup>52</sup> | Peer-reviewed publication | August 2020 | Cross-sectional | United States (unclear number of states) | For-profit, non-profit, and public | Y | 13,167 | For-profit: 9164 (69.6%)<br>Non-profit: 3265 (24.8 %)<br>Government: 737 (5.6%) | Outbreak (any cases), number of infections (dichotomised as >10%confirmed cases/beds or >20% confirmed + suspected cases/beds, or 10+ deaths), and deaths |
| (McGarry, Grabowski, & Barnett, 2020) <sup>33</sup> | Peer-reviewed publication | October 2020 | Cohort | United States (unclear number of states) | For-profit, non-profit, and public | Y | 15,035 | For-profit 10539 (70.1%)<br>Non-profit: 3518 (23.4%)<br>Government: 977 (6.5%) | PPE and staff shortage |
| (Gibson & Greene, 2020) <sup>38</sup> | Peer-reviewed publication | October 2020 | Cross-sectional | United States (unclear number of states) | For-profit, non-profit, and public | Y | 13,445 | For-profit: 9398 (69.9%)<br>Non-profit: 3200 (23.8%)<br>Government: 847 (6.3%) | Supply of PPE and staff shortage |
| (Rowan et al., 2020) <sup>29</sup> | Report by Mathematica | September 2020 | Cross-sectional | United States (Connecticut) | For-profit and non-profit | Y | 212 | For-profit: 176 (83%)<br>Non-profit: 36 (17%) | Outbreak (any cases), number of infections, and deaths per licensed bed |
| (Brown et al., 2020) <sup>31</sup> | Peer-reviewed publication (initially included as preprint) | November 2020 | Cross-sectional | Canada (Ontario) | For-profit, non-profit, and public (municipal). | N | 618 | For-profit: 358 (58%)<br>Non-profit: 159 (26%)<br>Municipal: 101 (16%) | Outbreaks (any cases), number of infections and deaths per 100 residents |
| (Burton et al., 2020) <sup>53</sup> | Peer-reviewed publication | October 2020 | Cross-sectional | Scotland | For-profit, non-profit, and public (local authority) | N | 189 | Private: 98 (52%)<br>Local authority: 54 (28%)<br>Non-profit: 37 (20%) | Outbreaks (any cases) |
| (Shen, 2020) <sup>54</sup> | Medrxiv preprint | September 2020 | Cross-sectional | United States (18 states) | For-profit and non-profit (including public) | Y | 6,132 | For-profit: 4476 (73%)<br>Non-profit: 1472 (24%)<br>Public: 184 (3%) | Number of deaths per 100 beds |
| (Chen, Chevalier, & Long, 2020) <sup>42</sup> | Peer-reviewed publication (initially included as preprint) | December 2020 | Cross-sectional | United States (22 states) | For-profit and non-profit | N | 6,337 | For-profit: 4689 (74%)<br>Non-profit: 1648 (26%) | Number of cases (inverse hyperbolic sine) |
| (Gandhi, Song, & Upadrashta, 2020) <sup>36</sup> | Preprint | October 2020 | Cross-sectional | United States (49 states) | Private equity, prior private equity, and for-profit | Y | 13,398 | Non-Private equity: 11,788 (88%)<br>Prior Private equity: 1,219 (9%)<br>Private equity: 391 (3%)<br>No info on for-profit | Number of confirmed and suspected infections, deaths, and PPE shortage |
| (Shallcross et al., 2020) <sup>41</sup> | Peer-reviewed publication (initially included as preprint) | February 2021 | Cross-sectional | England | For-profit and non-profit | Y | 5,126 | Profit: 4289 (83.7%)<br>Not-for-profit: 837 (16.3%) | Outbreaks (any cases) and number of infections (dichotomised as more than one third of staff or residents) |
| (Rolland et al., 2020) <sup>15</sup> | Peer-reviewed publication | July 2020 | Cross-sectional | France | For-profit, non-profit, and public | N | 124 | For-profit: 54 (44%)<br>Non-profit: 35 (28%)<br>Public: 35 (28%) | Outbreaks (any cases) |
| (Morciano, Stokes, Kontopanteli, Hall, & Turner, 2020) <sup>34</sup> | Medrxiv preprint | November 2020 | Cohort | England | For-profit and non-profit | Y | 15,524 | For-profit: 13397 (86.3%)<br>Non-profit: 2127 (14.7%) | Deaths (defined as at least one suspected or confirmed COVID-19 death) |
| (Gopal, Han, & Yaraghi, 2021) <sup>40</sup> | Peer-reviewed publication | January 2021 | Cross-sectional | United States (California) | For-profit and non-profit | N | 713 | For-profit: 613 (86%)<br>Non-profit: 100 (14%) | Outbreaks (any cases) and number of infections among residents and staff |

**Note:** Unless otherwise specified, outbreaks refer to the presence of at least one confirmed or suspected COVID-19 infection. Number of infections refer to the cumulative or relative number of confirmed or suspected COVID-19 infections. Deaths refer to the cumulative or relative mortality number unless otherwise specified. Only outcomes investigated in relation to ownership are included in this table.

Ownership was most commonly operationalised by comparing ‘for-profit’ (FP) and ‘non-profit’ (NP) care homes (17/29), usually with FPs as the reference category. Ten studies compared FPs and NPs (11/29), in which ‘non-profits’ also include public care homes, although this was not always explicitly described. Two studies focused on private equity (PE) providers in their operationalisation of ownership ^35,36^. Eighteen studies adjusted for the chain affiliation (CA) of care homes to investigate COVID-19 related outcomes.

The most investigated outcome was COVID-19 outbreak (16/29), followed by COVID-19 related mortality (13/29), and incidence of COVID-19 infections (13/29). Five studies investigated staff access to PPE and/or shortage of PPE ^33,35–38^. Five studies investigated COVID-19 related outcomes among care home staff ^36,39–41^. Most studies investigated multiple COVID-19 outcomes. Three studies were published after initially being included as preprints ^31,41,42^ and the results from the published versions are presented below.

#### Data sources and time coverage

Most of the included research merged multiple data sources on COVID-19 outcomes, information on the care homes, and area characteristics to construct their dataset. Less than 15% of included studies (4/29) collected primary data on the investigated COVID-19 outcomes. The majority of studies used data routinely reported by care homes to public health departments and other government-collected data. Almost a third of the included studies (9/29) used data from the Centers for Medicare & Medicaid Services (CMS), who required American nursing homes to report COVID-19 related data, including confirmed and suspected infections and deaths among residents and staff ^55^ from May 24. Providers were encouraged, but not required, to retrospectively self-report COVID-19 outcomes before this date. Table A2 in the supplementary material provides an overview of the data sources and the time period of the dependent variables across all included studies.

Eighteen studies reported on self-reported (confirmed and suspected cases) COVID-19 outcomes, whereas 19 studies only investigated confirmed COVID-19 outcomes. In one study, it was unclear if the investigated outcomes were confirmed or self-reported. All studies that investigated PPE and staffing shortage relied on self-reported outcomes. Most studies investigated COVID-19 outcomes collected in the March to July timespan with only two studies including data from later than September, 2020 ^54^. Most studies investigated outcomes covering a 1-2 months period although five studies investigated a period of less than two weeks ^36,37,39,43,45^. The findings presented below thus relate to the first wave of the pandemic.

### Risk of bias assessments

The RoB assessments are detailed in Table A3 in the supplementary material. The main concerns related to systematic missing data and selection bias in the included studies. For example, studies that investigated the characteristics of excluded observations from the CMS dataset (due to missing or incomplete data) found that excluded care homes were more likely to be FP and were also associated to many risk factors, such as the ethnicity and socio-economic status of residents (discussed in detail below) ^33,37–39^. This is a potentially serious limitation of the studies using this data (for the purpose of this review), as it suggests that poorly performing FPs may be systematically underrepresented in the sample, which may underestimate the observed effect of ownership on COVID-19 outcomes. Because of this limitation, all studies using this dataset were downgraded to (at least) moderate risk. For studies using public and government data, we assumed the risk of information bias to be low.

All assessments of confounding bias done as part of the RoB were based on consideration of factors known to exacerbate the effect of COVID-19 ^56–59^ and the performance of care homes for older people ^6,7^. Almost all studies adjusted for the size of the care homes (24/29), and characteristics on quality and staffing were also commonly included. However, only 5 studies adjusted their outcome(s) to the number of beds or residents ^29,31,35,42,54^. Nineteen studies adjusted for the ethnic composition and 15/29 included information about the socio-economic status of residents. Rurality and/or population density was included in 11/29 of the studies and local/community incidence of COVID-19 were controlled for in 17/29 of the studies. See Table A4 in the supplementary material for details on the direction of effect and model adjustment of all included studies.

### Direction of effects

Figure 2 displays the direction of effect for all included studies across different ownership and COVID-19 outcomes. Bar height indicates overall risk of bias (with taller bars indicating lower RoB) and colour denotes country context. See Figure A1 in the supplementary material for a harvest plot on the direction of effect across different data sources and Table 2 for details on the confidence of each finding.

**Table 2:**
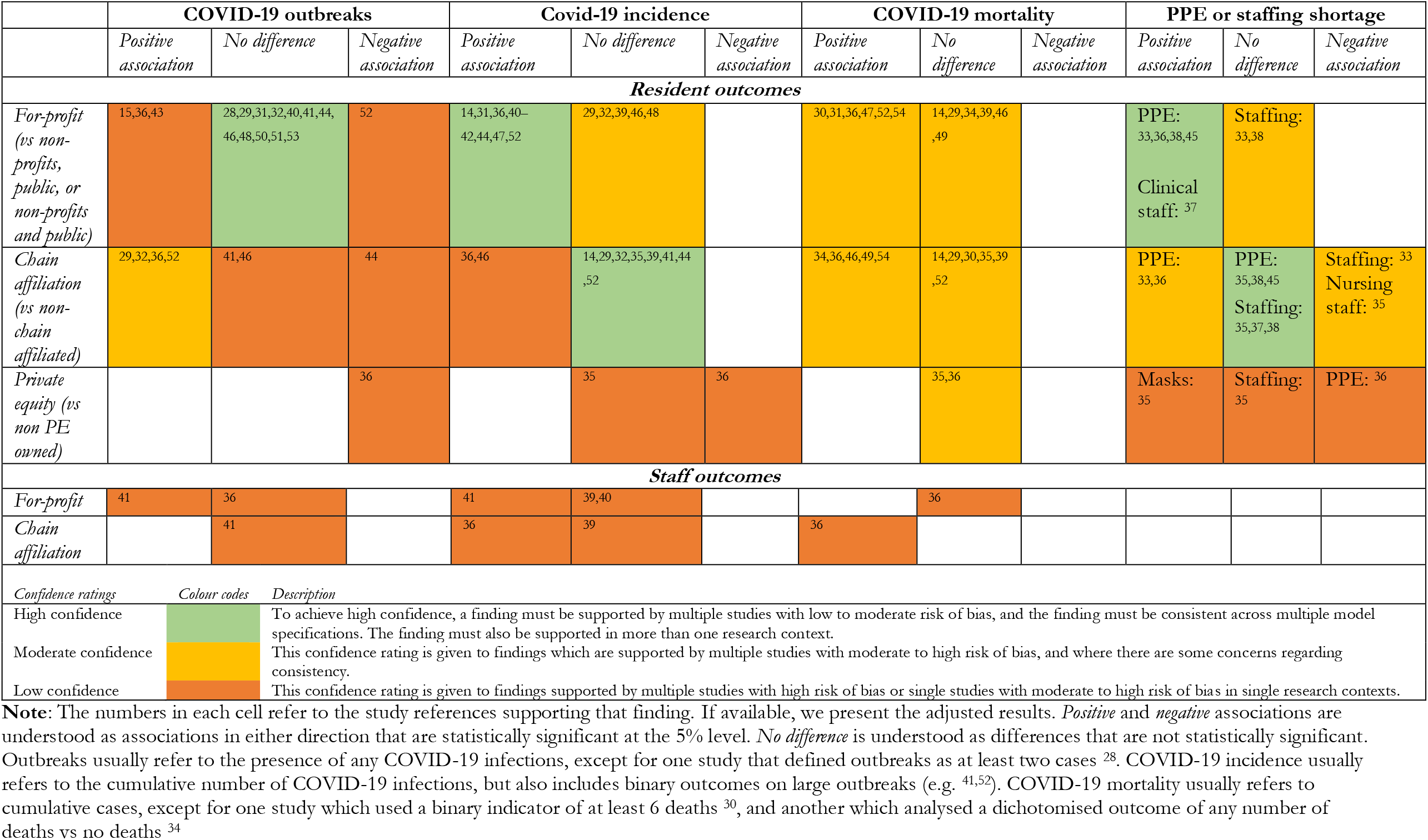
Overview of outcomes and confidence in findings.

**Figure 1:**
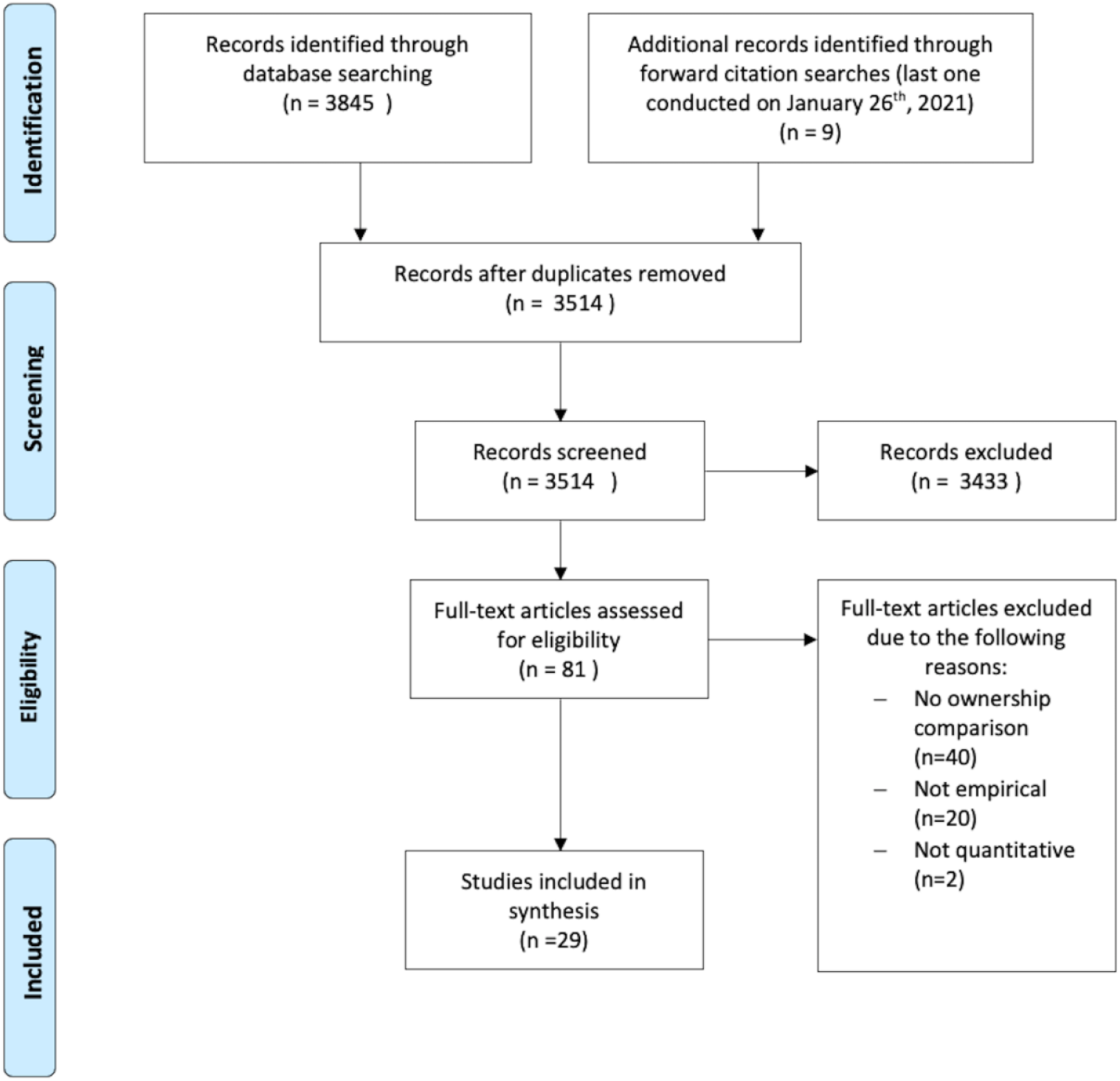
Prisma flow diagram.

**Figure 2:**
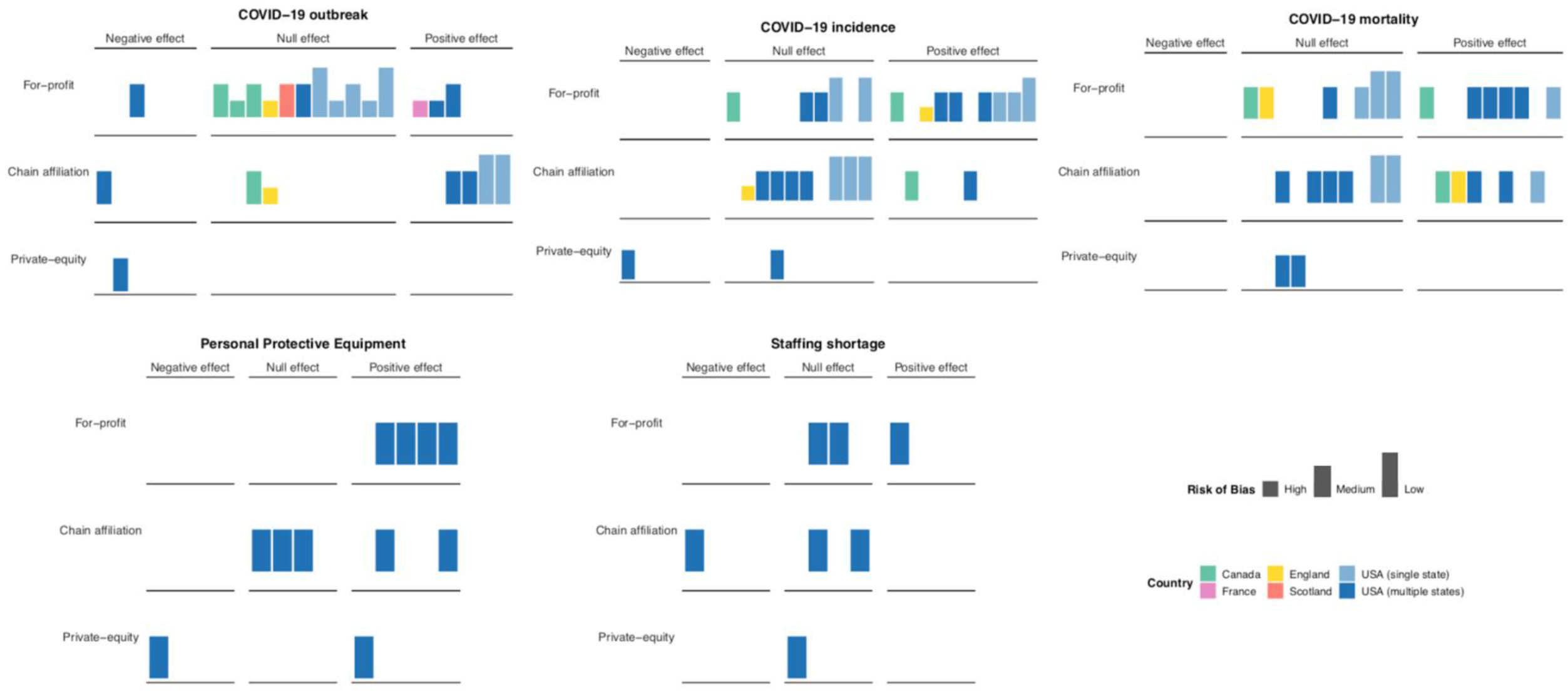
Harvest plot on the direction of effect across ownership, risk of bias, and study context. Bar height indicates overall risk of bias and colour denotes study context. *Positive* and *negative* effects are understood as associations in either direction that are statistically significant at the 5% level. Note that positive effects refer to elevated COVID-19 outcome values. *Null effect* is understood as differences that are not statistically significant at the 5% level. Outbreaks usually refer to the presence of any COVID-19 infections, except for one study that defined outbreaks as at least two cases ^28^. COVID-19 incidence usually refers to the cumulative number of COVID-19 infections, but also includes binary outcomes on large outbreaks (e.g. ^41,52^). COVID-19 mortality usually refers to cumulative cases, except for one study which used a binary indicator of at least 6 deaths ^30^, and another which analysed a dichotomised outcome of any number of deaths vs no deaths ^34^

#### Resident outcomes

In most studies investigating outbreaks (12/15) among residents, FP ownership was not associated with a higher risk of outbreak, suggesting that FP care homes were not more or less likely to have at least one infected resident. For-profit care homes were usually found to be associated with a higher number of cumulative COVID-19 infections. This direction of effect (i.e., positive effect) was coherent across multiple contexts, including the US, England, and Canada. No studies found FP ownership to be associated with fewer COVID-19 infections.

The evidence on COVID-19 mortality among residents and FP ownership was mixed. Six studies found higher rates of COVID-19 related deaths in FP care homes in Canada (Ontario ^31^), California ^47^, and across aggregated samples of US states ^30,36,52,54^, whereas six studies analysing data from Canada (Ontario ^46^), England ^34^, Connecticut ^14,29^, New York state ^49^, and a single study using aggregate data from multiple states ^39^ did not find a statistically significant association. However, the English study only investigated variation in the probability of having at least 1 COVID-19 related death and did not analyse variation in the cumulative numbers of deaths ^34^. More importantly, most of the US studies (4/5) which analysed cumulative state data (as opposed to data from single states) reported a statistically significant relationship between FP ownership and mortality ^30,36,52,54^. The only study using cumulative state data which did not identify statistically significant ownership variation analysed COVID-19 deaths for only one week (May 25 to May 31, 2020) ^39^. The opposing results regarding ownership in Canada (Ontario) are discussed in the below section. No studies found FP ownership to be associated with fewer COVID-19 deaths.

All studies that investigated PPE outcomes found FP ownership to be positively associated with insufficient access to or shortage of PPE ^33,36,38,45^. FP ownership was not consistently associated with staffing shortages.

Chain affiliated care homes were generally associated with a higher likelihood of COVID-19 outbreaks, but not with a higher incidence of infections. Five studies from Canada, England, and the US found CA ownership to be associated with a higher incidence of COVID-19 deaths, whereas 6 studies using both single and cumulative US state data did not identify any statistically significant variation. The one Canadian study that investigated this ownership category found CA care homes to be associated with more COVID-19 infections and deaths ^46^. The two English studies investigating this group did not identify any variation in outbreaks and incidence across chain ownership ^41^, but one found CA care homes to be associated with higher risk of at least one COVID-19 death among residents ^34^.

Care homes owned by private equity (PE) firms were not found to be consistently associated with worse outcomes than other ownership categories, and one study even found PE providers to be less likely to report PPE shortages and confirmed COVID-19 outbreaks ^36^.

#### Staff outcomes

Evidence on the relationship between ownership variation and risk of infection among staff was modest, with only four studies investigating this population. In England, FP ownership was associated with a higher incidence of infection among staff ^41^, but there was no difference for CA care homes. The three studies conducted in the US did not identify any statistically significant variation related to FP ownership, but one study found CA care homes to be correlated with a higher risk of infection and deaths among staff ^36^.

### Indirect ownership effects

Most studies reported the unadjusted prevalence of deaths and infections to be higher in FP care homes, but this effect was not always statistically significant when adjusting for covariates. This suggest that there are important mediating factors that influence the effect of ownership on COVID-19 outcomes. However, ownership was usually treated as a covariate for model adjustment, and the specific results relating to this variable were not often directly discussed and interpreted. For the studies which did discuss the effect of ownership on COVID-19 (16/29), the below mentioned mediating pathways were considered most important.

For-profit care homes were more likely to report PPE shortages ^33,35,38^, which will inevitably have influenced their ability to safeguard residents in the early wave of the pandemic. Some interpreted this association to mean that FP care homes were less willing to challenge the financial bottom-line by making additional investments, even during a pandemic ^14,33^. Yet private equity did not report PPE shortages ^35,36^, raising questions about whether the profit motive is the main driving factor.

Table 3 provides an overview of the key risk factors that were correlated to FP ownership and how adjusting for these factors influenced the observed impact of FP ownership on COVID-19 outcomes. There was evidence to suggest that the effect of ownership was sensitive to certain model adjustments. For example, one Canadian study reported that the effect of FP ownership was mediated by care home characteristics, which was operationalised as older care home design standards, number of residents, staff to bed ratio, and chain affiliation. When including these covariates, the effect of FP ownership on COVID-19 mortality and incidence lost statistical significance. However, it was not clear from the analysis which of the included care home characteristics accounted for this change. Notably, a study conducted by the same team analysing data on the identical sample (long-term care homes in Ontario), time period (March 29 to May 20, 2020), and outcomes (outbreaks, incidence, and mortalities) accounted for different covariates and came to different conclusions ^31,46^. Specifically, Stall et al ^46^ did not find FP ownership to be a statistically significant predictor of COVID-19 mortality and incidence in their fully adjusted models, whereas Brown et al ^31^, who adjusted for crowding instead of chain affiliation and outdated design standards, did report FP ownership to remain a statistically significant risk factor.

**Table 3:** Overview of risk factors associated to both FP ownership and COVID-19 outcomes.

| **COVID-19 risk factors correlated to FP ownership** | **Evidence supporting correlation between risk factor and FP ownership** | **Importance of risk factor** |
| --- | --- | --- |
| Chain affiliation | Research conducted in Connecticut <sup>14</sup>, Canada (Ontario) <sup>46</sup>, and England <sup>34</sup> found FP care homes to be more likely to be affiliated to a chain. | Of the 18 studies adjusting for chain ownership, 11 found FP ownership to exacerbate COVID-19 outcomes. |
| Resident ethnicity | A study using cumulative US state data found that FP care homes serve a higher proportion of ethnic minority residents <sup>39</sup>. | Of the 17 studies that adjusted for ethnicity, 12 found FP ownership to exacerbate COVID-19 outcomes |
| Older design standards | A study conducted in Canada (Ontario) found that outdated design standards were more prevalent in FP care homes <sup>46</sup>. | The one study adjusting for outdated design standards did not find FP ownership to be associated to inferior COVID-19 outcomes. |
| Crowding | A study conducted in Canada (Ontario) <sup>31</sup> found that FP care homes were more likely to be crowded. | The one study adjusting for crowding found that both crowding and FP ownership were statistically significant risk factors. |
| Staffing | A study conducted in California <sup>50</sup> found FP care homes to be associated with inferior staffing conditions. | Of the 18 studies adjusting for staffing conditions, 11 found FP ownership to exacerbate COVID-19 outcomes. |
| Quality | Research conducted in West Virginia <sup>28</sup> and California <sup>50</sup> found FP ownership to be correlated with poor quality ratings. | Of the 18 studies adjusting for quality, 10 found FP ownership to exacerbate COVID-19 outcomes. |
| Union membership | A study conducted in New York state found that FP care homes were more likely to be part of a union <sup>49</sup>. | The one study adjusting for union membership did not find FP ownership to be associated to inferior COVID-19 outcomes. |
| Care home size | Research conducted in Scotland <sup>53</sup> and Connecticut <sup>14</sup> reported FP ownership to be correlated to care home size. | Of the 25 studies adjusting for care home size, 14 found FP ownership to exacerbate COVID-19 outcomes. |

**Note:** This table only reports risk factors that were found to be significantly associated with FP ownership. Most studies which included the above mentioned risk factors as covariates did not report their correlation to FP ownership. See table A3 in the appendix for details on model adjustment and direction of effect.

FP care homes were also more likely to be larger (in terms of number of clients)^14^, to serve minority groups ^39^, to be more crowded ^31^, and to have lower quality ratings ^28,50^. This multicollinearity between ownership and important risk characteristics may account for a large part of the observed ownership effects. However, a substantial body of studies with low to moderate risk of bias found FP care home to perform worse during the pandemic, even when controlling for these characteristics. This consistent effect across country context and model specification suggests that there is a direct association between FP ownership and COVID-19 performance, independent of mediating variables, which is also consistent with previous research on care quality and FP ownership ^8,9^.

## Discussion

### Summary of findings

Our synthesis and critical appraisal of 29 studies suggests that FP ownership is not consistently associated with a higher probability of COVID-19 outbreaks. Yet, there is compelling evidence suggesting that the consequences of outbreaks, in terms of cumulative infections and deaths, may be exacerbated by FP ownership. The finding that FP providers were consistently associated with PPE shortages, may help to explain why these care homes suffered from higher rates of infections and deaths following a COVID-19 outbreak during the early stages of the pandemic. However, there are important risk factors that may contribute to mediating this relationship, as FP care homes were often found to be associated to other important risk factors such as crowdedness ^31^, client vulnerability ^39^, and inferior quality ratings ^28,50^.

Chain affiliated providers were often found to be correlated with higher risk of outbreak, but were not consistently associated with elevated numbers of infections. There was some evidence demonstrating a higher incidence of COVID-19 deaths among chain affiliated providers, particularly in Canada and England. Private-equity ownership was not consistently associated with inferior COVID-19 outcomes.

### Implications

The findings of this review highlight the importance of ownership in accounting for poor COVID-19 outcomes across care homes. It is known that the adult social care sector found itself exceedingly exposed in the beginning of the pandemic ^60^, in large part due to delayed government support and intervention, but also as a result of many years of political and financial neglect ^11^. With this review, we do not suggest that the challenges faced by care homes during the pandemic can (or should) be understood through the lens of ownership alone. It is clear that all care homes have faced severe challenges, which cannot be reduced to ownership. However, outsourcing to for-profit providers has become the status-quo in many care markets, often based on the rationale that open market competition will optimise the functioning of care homes. This claim has been extensively criticised and is not supported by empirical work ^6–9,61^. This review adds to this evidence base by systematically appraising and synthesising the available research on how the consequences of the COVID-19 pandemic in care homes has varied by ownership during the first wave of the pandemic. Although our results represent multiple national settings, most of the included research was conducted in the US due to the availability of the national CMS dataset. Efforts are currently being made in the UK to create a similar type of systematic, live, and linked dataset on care homes ^62^, which is an important endeavour if the consequences of this pandemic are to be understood and addressed going forward.

### Limitations

Our findings should be interpreted in light of certain caveats, most of which relate to the characteristics of the included studies. First, most studies were conducted in the US and Canada and the results thus primarily relate to North America. Second, the majority of the US studies relied on CMS data, whereas all the Canadian studies were conducted in Ontario using the same sample of long term care facilities, which means that there may be overlap in the analysed data across certain studies. Although most studies investigated different outcomes and time periods, two Canadian studies analysed data covering the same sample, outcomes, and time interval ^31,46^. Second, the body of included research was too heterogenous to be meaningfully meta-analysed and this version of the review thus represents a critical appraisal and narrative synthesis conducted in line with the SWiM and COSMOS-E guidance ^23,26^. Third, throughout our risk of bias assessments, we assumed that the reporting of COVID-19 outcomes was not systematically related to ownership. However, there is some suggestive evidence of a longer turnaround period for resident test results among for-profit providers ^63^, which if generally true, may bias the effect of FP ownership towards the null due to underreporting. Last, it is known that COVID-19 research is rapidly published ^19^, which may expose our results to publication bias for articles that have been fast tracked for reporting timely and significant outcomes. However, by not restricting our studies to peer-reviewed research, we were able to also consider evidence presented in preprints and government reports in our synthesis.

## Conclusion and future research

This review constitutes the first version of a living appraisal and synthesis of evidence on ownership variation across COVID-19 outcomes. It will be updated as new research becomes available, which may change the conclusion of our synthesis. Based on our synthesis of the available research, we find FP ownership to be a consistent and credible risk factor of higher cumulative COVID-19 infections and deaths in the first wave of the pandemic. Therefore, ownership and the characteristics associated with FP care home providers may present key regulatable factors that can be addressed to improve health outcomes in vulnerable populations and reduce health disparities. Going forward, we hope future research will incorporate other national contexts and clearly define their ownership categories of interest.

## Data Availability

Most of the data represented in available in the manuscript or in the supplementary material. Additional data is available upon request. Our protocol is available online on Prospero (CRD42020218673) and on OSF (osf.io/c8dq9/). The authors affirm that this manuscript is an honest, accurate, and transparent account of the study being reported; that no important aspects of the study have been omitted; and that any discrepancies from the study as planned have been explained.

## Contributors

ABM conceived the idea for the manuscript and designed the study protocol with feedback from BV and MDE. ABM and MDE double-screened and selected the included studies. ABM extracted and analysed all data with feedback from MDE and BV. AM validated all the extraction results. All authors contributed to quality assessing the included research in duplicate. MDE and ABM developed the visualisations for the paper. ABM wrote the manuscript with feedback from MDE, AM, and BV. The corresponding author attests that all listed authors meet authorship criteria and that no others meeting the criteria have been omitted.

## Funding

Anders Bach-Mortensen is supported by a research fellowship from the Carlsberg Foundation. The funder was not involved in the research process.

## Competing interests

None.

## Ethical approval

Not applicable.

## Data sharing

Most of the data represented is available in the manuscript or in the supplementary material. Additional data is available upon request. Our protocol is available online on Prospero (CRD42020218673) and on OSF (osf.io/c8dq9/). The authors affirm that this manuscript is an honest, accurate, and transparent account of the study being reported; that no important aspects of the study have been omitted; and that any discrepancies from the study as planned have been explained.

## Dissemination to participants and related patient and public communities

Our protocol is available online on Prospero (CRD42020218673) and on OSF (osf.io/c8dq9/).

## Supplementary Material

**Table A1:** Ownership categorisation and overlap.

| Ownership categories | Definition | Potential overlap |
| --- | --- | --- |
| For-profit | Private profit pursuing providers of social care. This usually includes both individually owned, partnerships, and corporate providers. | This term may include **chain affiliated** and **private equity owned** providers. **For-profits** and **non-profit** providers are sometimes categorised as **private** providers. |
| Non-profit | Private not-for-profit providers of social care. These providers are usually registered as charities. | This term may include **chain affiliated** providers. **For-profits** and **non-profit** providers are sometimes categorised as **private** providers. |
| Public | Care providers operated and organised by government bodies and employees. These providers are usually operated at the municipal, council, or local authority level. | **Public** and **non-profit** providers are often categorised as **not-for-profit** providers. |
| Private | Providers that are privately owned, including both for-profit and non-profit providers. | This may include **for-profit, non-profit, chain affiliated, and private equity** providers. |
| Multifacility chain affiliation | Providers that are part of a multi-location chain of social care facilities. | This may include both **for-profit, private equity, and non-profit** providers. |
| Private equity owned | Providers that have been acquisitioned by private equity firms. | This may include **for-profit** and **chain affiliated** providers. |

**Table A2:** Data sources and time period for COVID-19 outcomes.

| Study | Time period for COVID-19 outcomes | Data source | Confirmed or self-reported outcomes |
| --- | --- | --- | --- |
| <sup>1</sup> | March 17 to June 11, 2020 | West Virginia Department of Health and Human Resources | Confirmed |
| <sup>2</sup> | April 22 to April 29, 2020 | Cumulative state data | Self-report (confirmed and suspected) |
| <sup>3</sup> | As of May 11, 2020 | Cumulative state data | Self-report (confirmed and suspected) |
| <sup>4</sup> | May 17 to July 2, 2020 | CMS data | Self-report (confirmed and suspected) |
| <sup>5</sup> | As of April 16, 2020 | Connecticut Department of Health and Human Services | Confirmed |
| <sup>6</sup> | Week ending June 21, 2020 | CMS data | Self-report (confirmed and suspected) |
| <sup>7</sup> | March 29 to May 20, 2020 | Ontario Ministry of Health and Long-term Care | Confirmed |
| <sup>8</sup> | April 23 to June 2, 2020 | California Department of Public Health (California SNF's COVID-19 data) | Confirmed |
| <sup>9</sup> | As of June 30 2020 | CMS data | Self-report (confirmed and suspected) |
| <sup>10</sup> | As of May 31<sup>st</sup>, 2020 | New York State Department of Health | Confirmed |
| <sup>11</sup> | March to May 4, 2020 | California and Los Angeles Departments of Public Health | Self-report (confirmed and suspected) |
| <sup>12</sup> | Connecticut: As of April 16<br>New York: As of April 15<br>New Jersey: As of April 20 | State data from Connecticut, New Jersey, and New York. | Unclear |
| <sup>13</sup> | Weekly reports on April 28, May 14, and June 17 – all recorded from April 15, 2020 | Ohio Department of Health | Confirmed |
| <sup>14</sup> | May 25 to May 31, 2020 | CMS data | Self-report (confirmed and suspected) |
| <sup>15</sup> | March 3 to April 11, 2020 | Ontario Ministry of Health and Long-term Care | Confirmed |
| <sup>16</sup> | The week of May 31, 2020 | CMS data | Self-report (confirmed and suspected) |
| <sup>17</sup> | As of June 25, 2020 | CMS data | Self-report (confirmed and suspected) |
| <sup>18</sup> | May 18 to June 14, 2020 (study period 1) and June 24 to July 19, 2020 (study period 2). | CMS data | Self-report (confirmed and suspected) |
| <sup>19</sup> | Week ending June 21, to the week ending July 19, 2020 | CMS data | Self-report (confirmed and suspected) |
| <sup>20</sup> | As of July 22, 2020 | Connecticut Department of Health and Human Services | Self-report (confirmed and suspected) |
| <sup>21</sup> | March 29 to May 20, 2020 | Ontario Ministry of Health and Long-term Care | Self-report (confirmed and suspected) |
| <sup>22</sup> | March 1 to August 2, 2020 | Care Inspectorate | Confirmed |
| <sup>23</sup> | May 6 to 7 October, 2020 | Cumulative state data | Self-report (confirmed and suspected) |
| <sup>24</sup> | As of May 31, 2020 | Cumulative state data | Self-report (confirmed and suspected) |
| <sup>25</sup> | Week ending May 24, 2020 | CMS data | Self-report (confirmed and suspected) |
| <sup>26</sup> | April 30 to 13 June, 2020 | Unique data collected for the study (conducted by Ipsos MORI) | Confirmed |
| <sup>27</sup> | March 23rd to May 6<sup>th</sup>, 2020 | Survey data collected for this paper | Self-report (confirmed and suspected) |
| <sup>28</sup> | March 4 to August 7, 2020 | Care Quality Commission | Self-report (confirmed and suspected) |
| <sup>29</sup> | As of 1<sup>st</sup> May, 2020 | California Department of Public Health | Confirmed |

CMS: Centers for Medicare and Medicaid services, SNF: Skilled Nursing Facility data.

**Table A3:** Risk of bias assessments using COSMOS-E guidance.

| **Study** | **Risk of bias** |  |  |
| --- | --- | --- | --- |
|  | *Confounding bias* | *Selection bias* | *Information bias* |
| (Bui et al., 2020) <sup>25</sup> | High | Low | Low |
| (Chatterjee, Kelly, Qi, & Werner, 2020) <sup>40</sup> | High | Low | Low |
| (Abrams, Loomer, Gandhi, & Grabowski, 2020) <sup>41</sup> | Moderate | Moderate | Moderate |
| (Braun et al., 2020) <sup>32</sup> | Low | Moderate | Low |
| (Yue Li, Temkin-Greener, Gao, & Cai, 2020) <sup>14</sup> | Low | Low | Low |
| (Yumeng Li, Fang, & He, 2020) <sup>42</sup> | Low | Moderate | Low |
| (Stall, Jones, Brown, Rochon, & Costa, 2020) <sup>43</sup> | Moderate | Moderate | Low |
| (He, Li, & Fang, 2020) <sup>44</sup> | Moderate | Low | Low |
| (Sugg et al., 2021) <sup>45</sup> | Moderate | Moderate | Low |
| (Dean, Venkataramani, & Kimmel, 2020) <sup>46</sup> | Low | Moderate | Low |
| (Harrington et al., 2020) <sup>47</sup> | Moderate | High | Low |
| (Unruh, Yun, Zhang, Braun, & Jung, 2020) <sup>27</sup> | Moderate | Low | Low |
| (Bowblis & Applebaum, 2020) <sup>29</sup> | Low | Low | Low |
| (Yue Li, Cen, Cai, & Temkin-Greener, 2020) <sup>36</sup> | Low | Moderate | Low |
| (Fisman, Bogoch, Lapointe-Shaw, McCready, & Tuite, 2020) <sup>48</sup> | High | Low | Low |
| (Xu, Intrator, & Bowblis, 2020) <sup>34</sup> | Low | Moderate | Low |
| (Gorges & Konetzka, 2020) <sup>49</sup> | Low | Moderate | Low |
| (McGarry, Grabowski, & Barnett, 2020) <sup>30</sup> | Low | Moderate | Low |
| (Gibson & Greene, 2020) <sup>35</sup> | Low | Moderate | Low |
| (Rowan et al., 2020) <sup>26</sup> | Low | Low | Low |
| (Brown et al., 2020) <sup>28</sup> | Moderate | Moderate | Low |
| (Burton et al., 2020) <sup>50</sup> | Moderate | Low | Low |
| (Shen, 2020) <sup>51</sup> | Low | Moderate | Low |
| (Chen, Chevalier, & Long, 2020) <sup>39</sup> | Low | Moderate | Low |
| (Gandhi, Song, & Upadrashta, 2020) <sup>33</sup> | Low | Moderate | Low |
| (Shallcross et al., 2020) <sup>38</sup> | Low | High | Moderate |
| (Rolland et al., 2020) <sup>15</sup> | High | Moderate | Moderate |
| (Morciano, Stokes, Kontopantelis, Hall, & Turner, 2020) <sup>31</sup> | Moderate | Low | Low |
| (Gopal, Han, & Yaraghi, 2021) <sup>37</sup> | Moderate | Moderate | Moderate |

**Table A4:**
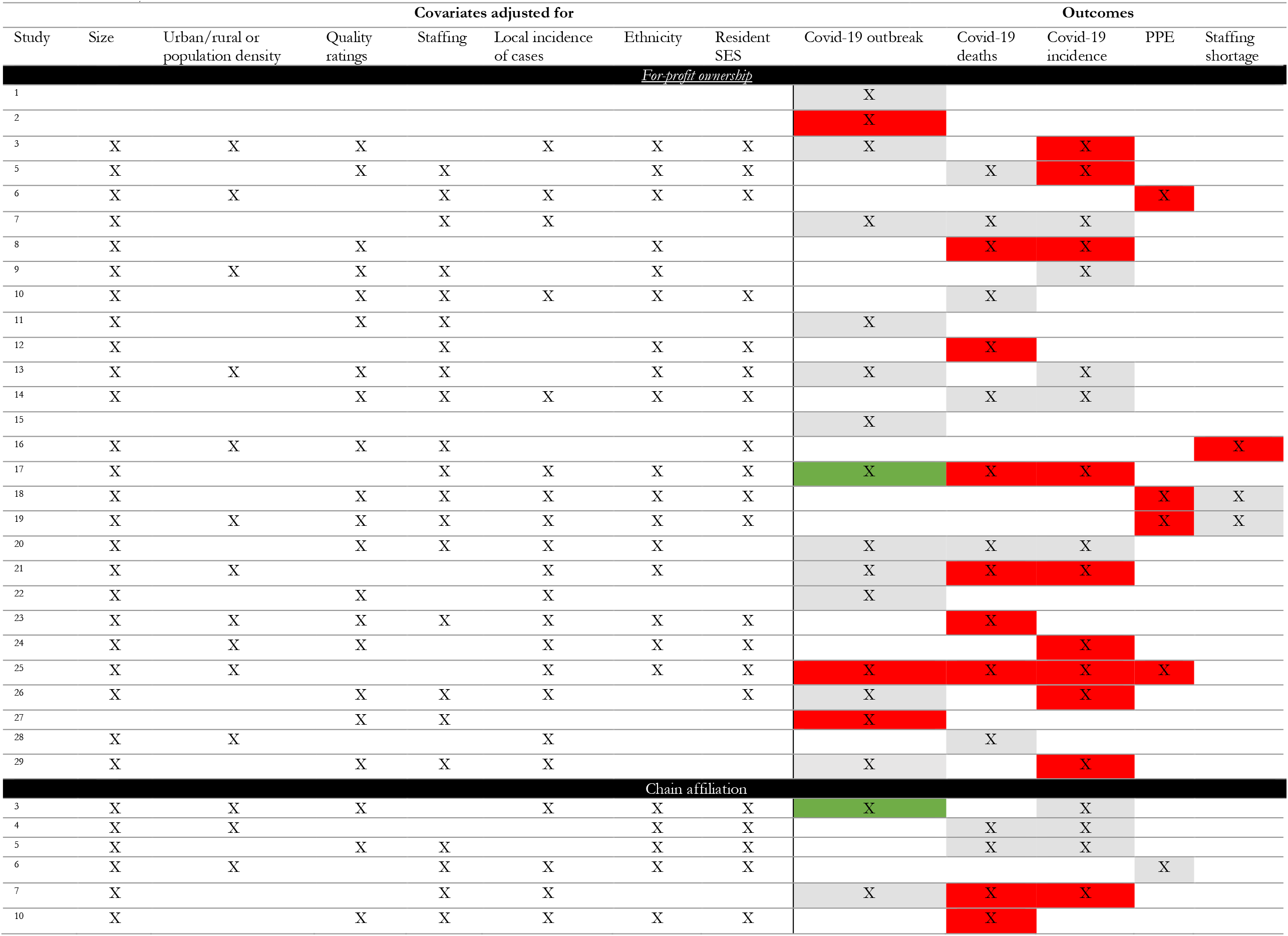
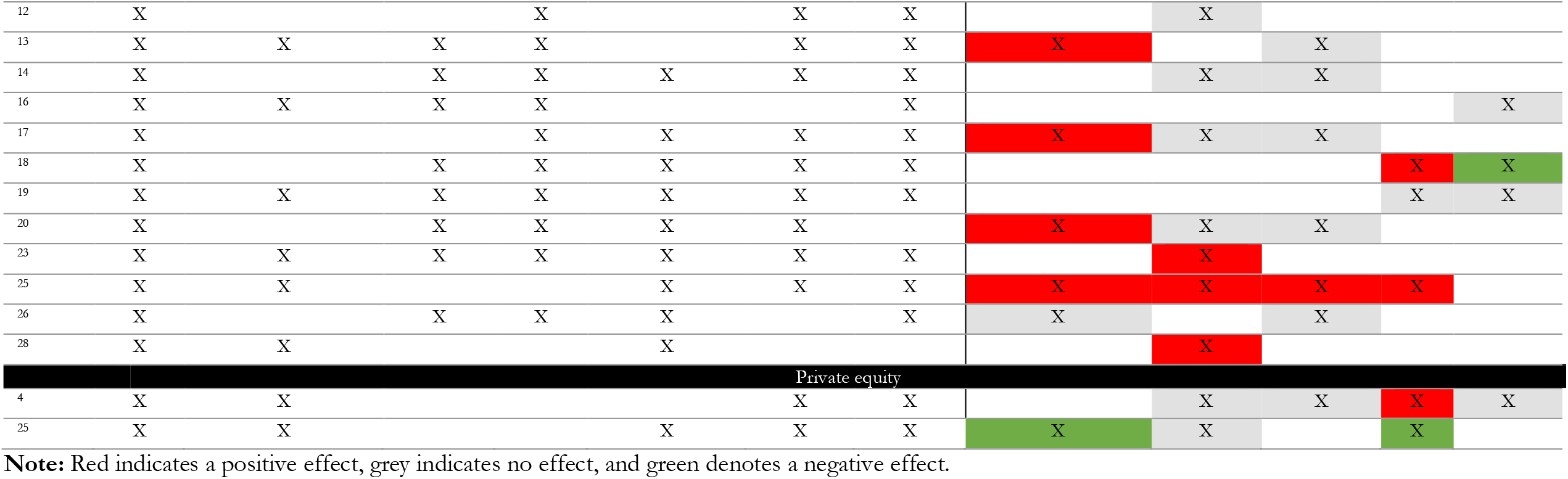
Adjustments and direction of effect.

**Figure A1:**
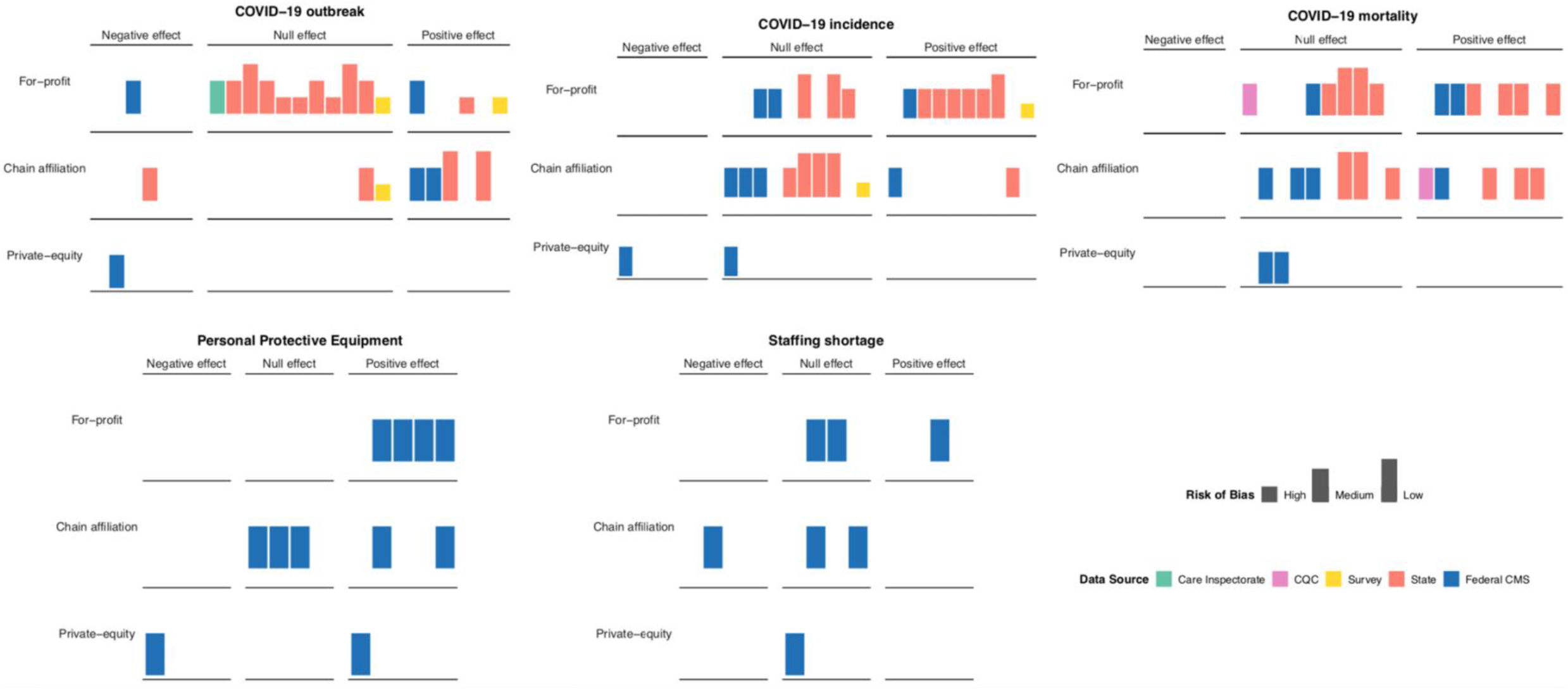
Harvest plot on the direction of effect across ownership, risk of bias, and data source. Bar height indicates overall risk of bias and colour denotes data source context. *Positive* and *negative* effects are understood as associations in either directions that are statistically significant at the 5% level. Note that positive effects refer to elevated COVID-19 outcome values. *Null effect* is understood as differences that are not statistically significant at the 5% level. Outbreaks usually refer to the presence of any COVID-19 infections, except for one study that defined outbreaks as at least two cases ^1^. COVID-19 incidence usually refers to the cumulative number of COVID-19 infections, but also includes binary outcomes on large outbreaks (e.g. ^17,26^). COVID-19 mortality usually refers to cumulative cases, except for one study which used a binary indicator of at least 6 deaths ^12^, and another which analysed a dichotomised outcome of any number of deaths vs no deaths ^28^. CQC: Care Quality Commission.

## PRISMA 2009 Checklist

| Section/topic | # | Checklist item | Reported on page # |
| --- | --- | --- | --- |
| **TITLE** |  |  |  |
| Title | 1 | Identify the report as a systematic review, meta-analysis, or both. | Title |
| **ABSTRACT** |  |  |  |
| Structured summary | 2 | Provide a structured summary including, as applicable: background; objectives; data sources; study eligibility criteria, participants, and interventions; study appraisal and synthesis methods; results; limitations; conclusions and implications of key findings; systematic review registration number. | 2 |
| **INTRODUCTION** |  |  |  |
| Rationale | 3 | Describe the rationale for the review in the context of what is already known. | 3 |
| Objectives | 4 | Provide an explicit statement of questions being addressed with reference to participants, interventions, comparisons, outcomes, and study design (PICOS). | 3 |
| **METHODS** |  |  |  |
| Protocol and registration | 5 | Indicate if a review protocol exists, if and where it can be accessed (e.g., Web address), and, if available, provide registration information including registration number. | 3 |
| Eligibility criteria | 6 | Specify study characteristics (e.g., PICOS, length of follow-up) and report characteristics (e.g., years considered, language, publication status) used as criteria for eligibility, giving rationale. | 4-5 |
| Information sources | 7 | Describe all information sources (e.g., databases with dates of coverage, contact with study authors to identify additional studies) in the search and date last searched. | 5 |
| Search | 8 | Present full electronic search strategy for at least one database, including any limits used, such that it could be repeated. | Supplementary material |
| Study selection | 9 | State the process for selecting studies (i.e., screening, eligibility, included in systematic review, and, if applicable, included in the meta-analysis). | 4-5 |
| Data collection process | 10 | Describe method of data extraction from reports (e.g., piloted forms, independently, in duplicate) and any processes for obtaining and confirming data from investigators. | 5 |
| Data items | 11 | List and define all variables for which data were sought (e.g., PICOS, funding sources) and any assumptions and simplifications made. | 5 |
| Risk of bias in individual studies | 12 | Describe methods used for assessing risk of bias of individual studies (including specification of whether this was done at the study or outcome level), and how this information is to be used in any data synthesis. | 5 |
| Summary measures | 13 | State the principal summary measures (e.g., risk ratio, difference in means). | NA |
| Synthesis of results | 14 | Describe the methods of handling data and combining results of studies, if done, including measures of consistency (e.g., $I^2$) for each meta-analysis. | 5-6 |
| Risk of bias across studies | 15 | Specify any assessment of risk of bias that may affect the cumulative evidence (e.g., publication bias, selective reporting within studies). | 5 |
| Additional analyses | 16 | Describe methods of additional analyses (e.g., sensitivity or subgroup analyses, meta-regression), if done, indicating which were pre-specified. | 5-6 |
| **RESULTS** |  |  |  |
| Study selection | 17 | Give numbers of studies screened, assessed for eligibility, and included in the review, with reasons for exclusions at each stage, ideally with a flow diagram. | 6-10, Figure 1 |
| Study characteristics | 18 | For each study, present characteristics for which data were extracted (e.g., study size, PICOS, follow-up period) and provide the citations. | Table 1 |
| Risk of bias within studies | 19 | Present data on risk of bias of each study and, if available, any outcome level assessment (see item 12). | Table A3 |
| Results of individual studies | 20 | For all outcomes considered (benefits or harms), present, for each study: (a) simple summary data for each intervention group (b) effect estimates and confidence intervals, ideally with a forest plot. | Figure 2 |
| Synthesis of results | 21 | Present results of each meta-analysis done, including confidence intervals and measures of consistency. | Figure 2, Figure A1, Table 2 |
| Risk of bias across studies | 22 | Present results of any assessment of risk of bias across studies (see Item 15). | 10, Table A2 |
| Additional analysis | 23 | Give results of additional analyses, if done (e.g., sensitivity or subgroup analyses, meta-regression [see Item 16]). | 14-15 |
| **DISCUSSION** |  |  |  |
| Summary of evidence | 24 | Summarize the main findings including the strength of evidence for each main outcome; consider their relevance to key groups (e.g., healthcare providers, users, and policy makers). | 15 |
| Limitations | 25 | Discuss limitations at study and outcome level (e.g., risk of bias), and at review-level (e.g., incomplete retrieval of identified research, reporting bias). | 16 |
| Conclusions | 26 | Provide a general interpretation of the results in the context of other evidence, and implications for future research. | 15-16 |
| **FUNDING** |  |  |  |
| Funding | 27 | Describe sources of funding for the systematic review and other support (e.g., supply of data); role of funders for the systematic review. | 13 |

From: Moher D, Liberati A, Tetzlaff J, Altman DG, The PRISMA Group (2009). Preferred Reporting Items for Systematic Reviews and Meta-Analyses: The PRISMA Statement. PLoS Med 6(7): e1000097. doi:10.1371/journal.pmed1000097
For more information, visit: www.prisma-statement.org.

## Notes

### Competing Interest Statement

The authors have declared no competing interest.

### Clinical Protocols

https://www.crd.york.ac.uk/prospero/display_record.php?RecordID=218673

https://osf.io/c8dq9/

### Summary of Updates

This version constitutes the latest iteration of our living systematic review.

