## Supplementary material for "Ownership and COVID-19 in care homes for older people: A living systematic review of outbreaks, infections, and mortalities" (from another version of this preprint: version 1, 2021-01-29)

**Table A1:** Data sources and time period for COVID-19 outcomes.

| Study | Time period for COVID-19 outcomes | Data source |
|---|---|---|
| 1 | March 17 to June 11, 2020 | West Virginia Department of Health and Human Resources |
| 2 | April 22 to April 29, 2020 | Cumulative state data |
| 3 | As of May 11, 2020 | Cumulative state data |
| 4 | May 17 to July 2, 2020 | CMS data |
| 5 | As of April 16, 2020 | Connecticut Department of Health and Human Services |
| 6 | Week ending June 21, 2020 | CMS data |
| 7 | March 29 to May 20, 2020 | Ontario Ministry of Long-term Care |
| 8 | April 23 to June 2, 2020 | California Department of Public Health (California SNFs COVID-19 data) |
| 9 | As of June 30 2020 | CMS data |
| 10 | As of May 31st, 2020 | New York State Department of Health |
| 11 | March to May 4, 2020 | California and Los Angeles Departments of Public Health |
| 12 | Connecticut: As of April 16<br>New York: As of April 15<br>New Jersey: As of April 20 | State data from Connecticut, New Jersey, and New York. |
| 13 | Weekly reports on April 28, May 14, and June 17 – all recorded from April 15, 2020 | Ohio Department of Health |
| 14 | May 25 to May 31, 2020 | CMS data |
| 15 | March 3 to April 11, 2020 | Ontario Ministry of Health and Long-term Care |
| 16 | The week of May 31, 2020 | CMS data |
| 17 | As of June 25, 2020 | CMS data |
| 18 | May 18 to June 14, 2020 (study period 1) and June 24 to July 19, 2020 (study period 2). | CMS data |
| 19 | Week ending June 21, to the week ending July 19, 2020 | CMS data |
| 20 | As of July 22, 2020 | Connecticut Department of Health and Human Services |

| | | |
|---|---|---|
| 21 | March 29 to May 20, 2020 | Ontario Ministry of Health and Long-term Care |
| 22 | March 1 to August 2, 2020 | Care Inspectorate |
| 23 | May 6 to 7 October, 2020 | Cumulative state data |
| 24 | As of May 31, 2020 | Cumulative state data |
| 25 | Week ending May 24, 2020 | CMS data |
| 26 | April 30 to 13 June, 2020 | Unique data collected for the study (conducted by Ipsos MORI) |
| 27 | March 23rd to May 6th, 2020 | Survey data collected for this paper |
| 28 | March 4 to August 7, 2020 | Data provided by the Care Quality Commission |

CMS: Centers for Medicare and Medicaid services, SNF: Skilled Nursing Facility data.

**Table A2:** Risk of bias assessments using COSMOS-E guidance.

| Study | Risk of bias | | |
|---|---|---|---|
| | ***Confounding bias*** | ***Selection bias*** | ***Information bias*** |
| (Bui et al., 2020) [1] | High | Low | Low |
| (Chatterjee, Kelly, Qi, & Werner, 2020) [2] | High | Low | Low |
| (Abrams, Loomer, Gandhi, & Grabowski, 2020) [3] | Moderate | Moderate | Moderate |
| (Braun et al., 2020) [4] | Low | Moderate | Low |
| (Yue Li, Temkin-Greener, Gao, & Cai, 2020) [5] | Low | Low | Low |
| (Yumeng Li, Fang, & He, 2020) [6] | Low | Moderate | Low |
| (Stall, Jones, Brown, Rochon, & Costa, 2020) [7] | Moderate | Moderate | Low |
| (He, Li, & Fang, 2020) [8] | Moderate | Low | Low |
| (Sugg et al., 2021) [9] | Moderate | Moderate | Low |
| (Dean, Venkataramani, & Kimmel, 2020) [10] | Low | Moderate | Low |
| (Harrington et al., 2020) [11] | Moderate | High | Low |
| (Unruh, Yun, Zhang, Braun, & Jung, 2020) [12] | Moderate | Low | Low |
| (Bowblis & Applebaum, 2020) [13] | Low | Low | Low |
| (Yue Li, Cen, Cai, & Temkin-Greener, 2020) [14] | Low | Moderate | Low |
| (Fisman, Bogoch, Lapointe-Shaw, McCready, & Tuite, 2020) [15] | High | Low | Low |
| (Xu, Intrator, & Bowblis, 2020) [16] | Low | Moderate | Low |
| (Gorges & Konetzka, 2020) [17] | Low | Moderate | Low |
| (McGarry, Grabowski, & Barnett, 2020) [18] | Low | Moderate | Low |
| (Gibson & Greene, 2020) [19] | Low | Moderate | Low |
| (Rowan et al., 2020) [20] | Low | Low | Low |
| (Brown et al., 2020) [21] | Moderate | Low | Low |
| (Burton et al., 2020) [22] | Moderate | Low | Low |
| (Shen, 2020) [23] | Low | Moderate | Low |

| | | | |
|---|---|---|---|
| (Chen, Chevalier, & Long, 2020) [24] | Low | Moderate | Low |
| (Gandhi, Song, & Upadrashta, 2020) [25] | Low | Moderate | Low |
| (Shallcross et al., 2020) [26] | Low | High | Moderate |
| (Rolland et al., 2020) [27] | High | Moderate | Moderate |
| (Morciano, Stokes, Kontopantelis, Hall, & Turner, 2020) [28] | Moderate | Low | Low |

**Table A3:** Adjustments and direction of effect.

| | Covariates adjusted for | | | | | | | Outcomes | | | | |
|---|---|---|---|---|---|---|---|---|---|---|---|---|
| Study | Size | Urban/rural or population density | Quality ratings | Staffing | Local incidence of cases | Ethnicity | Resident SES | Covid-19 outbreak | Covid-19 deaths | Covid-19 incidence | PPE | Staffing shortage |
| ***For-profit ownership*** | | | | | | | | | | | | |
| 1 | | | | | | | | X | | | | |
| 2 | | | | | | | | X | | | | |
| 3 | X | X | X | | X | X | X | X | | X | | |
| 5 | X | | X | X | | X | X | | X | X | | |
| 6 | X | X | | X | X | X | X | | | | X | |
| 7 | | | | | X | | | X | X | X | | |
| 8 | X | | X | | | X | | | X | X | | |
| 9 | X | X | X | X | | X | | X | | X | | |
| 10 | X | | X | X | X | X | X | | X | | | |
| 11 | X | | X | X | | | | X | | | | |
| 12 | X | | | X | | X | X | | X | | | |
| 13 | X | X | X | X | | X | X | X | | X | | |
| 14 | X | | X | X | X | X | X | | X | X | | |
| 15 | | | | | | | | X | | | | |
| 16 | X | X | X | X | | | X | | | | | X |
| 17 | X | | | X | X | X | X | X | X | X | | |
| 18 | X | | X | X | X | X | X | | | | X | X |
| 19 | X | X | X | X | X | X | X | | | | X | X |
| 20 | X | | X | X | X | X | | X | X | X | | |
| 21 | X | X | | | X | X | | X | X | X | | |
| 22 | X | | X | | X | | | X | | | | |
| 23 | X | X | X | X | X | X | X | | X | | | |
| 24 | X | X | X | | X | X | X | | | X | | |
| 25 | X | X | | | X | X | X | X | X | | X | |
| 26 | X | | X | X | X | | X | X | | X | | |
| 27 | | | X | X | | | | X | | | | |
| 28 | X | X | | | X | | | | X | | | |
| **Chain affiliation** | | | | | | | | | | | | |
| 3 | X | X | X | | X | X | X | X | | X | | |
| 5 | X | | X | X | | X | X | | X | X | | |
| 6 | X | X | | X | X | X | X | | | | X | |
| 7 | X | | | X | X | | | X | X | X | | |
| 10 | X | | X | X | X | X | X | | X | | | |
| 12 | X | | | X | | X | X | | X | | | |

| | | | | | | | | | | | | |
|---|---|---|---|---|---|---|---|---|---|---|---|---|
| 13 | X | X | X | X | | X | X | X | | X | | |
| 14 | X | | X | X | X | X | X | | X | X | | |
| 16 | X | X | X | X | | | X | | | | | X |
| 17 | X | | | X | X | X | X | X | X | X | | |
| 18 | X | | X | X | X | X | X | | | | X | X |
| 19 | X | X | X | X | X | X | X | | | | X | X |
| 20 | X | | X | X | X | X | | X | X | X | | |
| 23 | X | X | X | X | X | X | X | | X | | | |
| 25 | X | X | | | X | X | X | X | X | | X | |
| 26 | X | | X | X | X | | X | X | | X | | |
| 28 | X | X | | | X | | | | X | | | |
| **Private equity** | | | | | | | | | | | | |
| 4 | X | X | | | | X | X | | X | X | X | X |
| 25 | X | X | | | X | X | X | X | X | | X | |

**Note:** Red indicates a positive effect, grey indicates no effect, and green denotes a negative effect.

**Table A4:** Overview of outcomes and confidence in findings.

| Type of facility | COVID-19 outbreaks | | | Covid-19 incidence | | | COVID-19 mortality | | | PPE or staffing shortage | | |
|---|---|---|---|---|---|---|---|---|---|---|---|---|
| | *Positive association* | *No difference* | *Negative association* | *Positive association* | *No difference* | *Negative association* | *Positive association* | *No difference* | *Negative association* | *Positive association* | *No difference* | *Negative association* |
| ***Resident outcomes*** | | | | | | | | | | | | |
| *For-profit (vs non-profits or non-profits and government)* | 2,25,27 | 1,3,7,9,11,13,15,20–22,26 | 17 | 3,5,7,8,17,21,24,26 | 9,13,14,20 | | 7,8,12,17,21,23,25 | 5,10,14,20,28 | | PPE: 6,18,19,25 Clinical staff: [16] | Staffing: 18,19 | |
| *Chain affiliation (vs non-chain affiliated)* | 13,17,20,25 | 7,26 | 3 | 7 | 3–5,13,14,17,20,26 | | 7,10,23,25,28 | 4,5,12,14,17,20 | | PPE: 18,25 | PPE: 4,6,19 Staffing: 4,16,19 | Staffing: [18] Nursing staff: [4] |
| *Private equity (vs non PE owned)* | | | 25 | 4 | 4 | | | 4,25 | | Masks: [4] | Staffing: 4 | PPE: [25] |
| ***Staff outcomes*** | | | | | | | | | | | | |
| *For-profit* | 26 | 25 | | | 14 | | | 25 | | | | |
| *Chain affiliation* | | 26 | | 25 | 14 | | 25 | | | | | |

| *Confidence ratings* | *Colour codes* | *Description* |
|---|---|---|
| High confidence | | To achieve high confidence, a finding must be supported by multiple studies with low to moderate risk of bias, and the finding must be consistent across multiple model specifications. The finding must also be supported in more than one research context. |
| Moderate confidence | | This confidence rating is given to findings which are supported by multiple studies with moderate to high risk of bias, and where there are some concerns regarding consistency. |
| Low confidence | | This confidence rating is given to findings supported by multiple studies with high risk of bias or single studies with moderate to high risk of bias in single research contexts. |

**Note**: The numbers in each cell refer to the study references supporting that finding. If available, we present the adjusted results. *Positive* and *negative* associations are understood as associations in either directions that are statistically significant at the 5% level. *No difference* is understood as differences that are not statistically significant. Outbreaks usually refer to the presence of any COVID-19 infections, except for one study that defined outbreaks as at least two cases [1]. COVID-19 incidence usually refers to the cumulative number of COVID-19 infections, but also includes binary outcomes on large outbreaks (e.g. [17,26]). COVID-19 mortality usually refers to cumulative cases, except for one study which used a binary indicator of at least 6 deaths [12], and another which analysed a dichotomised outcome of any number of deaths vs no deaths [28].

### Figure Legends for supplementary material

**Figure A1:** Harvest plot on the direction of effect across ownership, risk of bias, and data source. Bar height indicates overall risk of bias and colour denotes data source context. *Positive* and *negative* effects are understood as associations in either directions that are statistically significant at the 5% level. Note that positive effects refer to elevated COVID-19 outcome values. *Null effect* is understood as differences that are not statistically significant at the 5% level. Outbreaks usually refer to the presence of any COVID-19 infections, except for one study that defined outbreaks as at least two cases [1]. COVID-19 incidence usually refers to the cumulative number of COVID-19 infections, but also includes binary outcomes on large outbreaks (e.g. [17,26]). COVID-19 mortality usually refers to cumulative cases, except for one study which used a binary indicator of at least 6 deaths [12], and another which analysed a dichotomised outcome of any number of deaths vs no deaths [28]. CQC: Care Quality Commission.

**Figure A2:** Directed acyclic graph of COVID-19 outcomes. This figure represent the hypothesised relationship between COVID-19 outcomes.

**Figure A3**: Directed acyclic graph of ownership, key risk factors, and COVID-19 outcomes. This figure represent the hypothesised relationships between COVID-19 outcomes, key risk factors, and ownership. The green lines indicate the direct effect of ownership on COVID-19 outcomes. The bold lines indicate mediating and other causal pathways on COVID-19 outcomes.

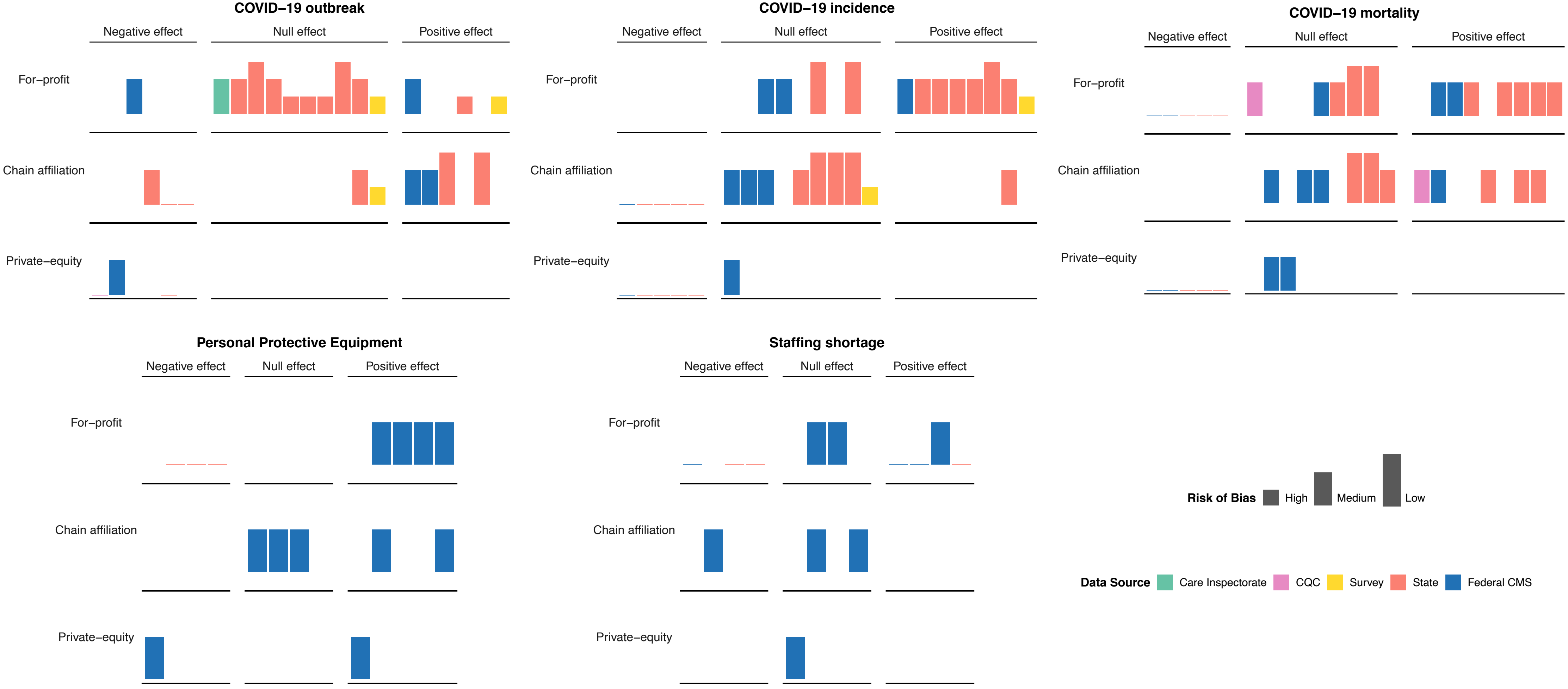
COVID–19 outbreak
COVID–19 incidence
COVID–19 mortality
Negative effect
Null effect
Positive effect
For–profit
Chain affiliation
Private–equity
Personal Protective Equipment
Staffing shortage
Risk of Bias
High
Medium
Low
Data Source
Care Inspectorate
CQC
Survey
State
Federal CMS

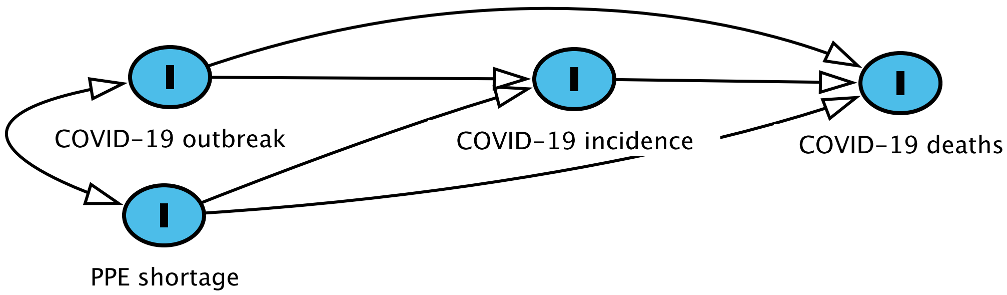
COVID-19 outbreak
COVID-19 incidence
COVID-19 deaths
PPE shortage

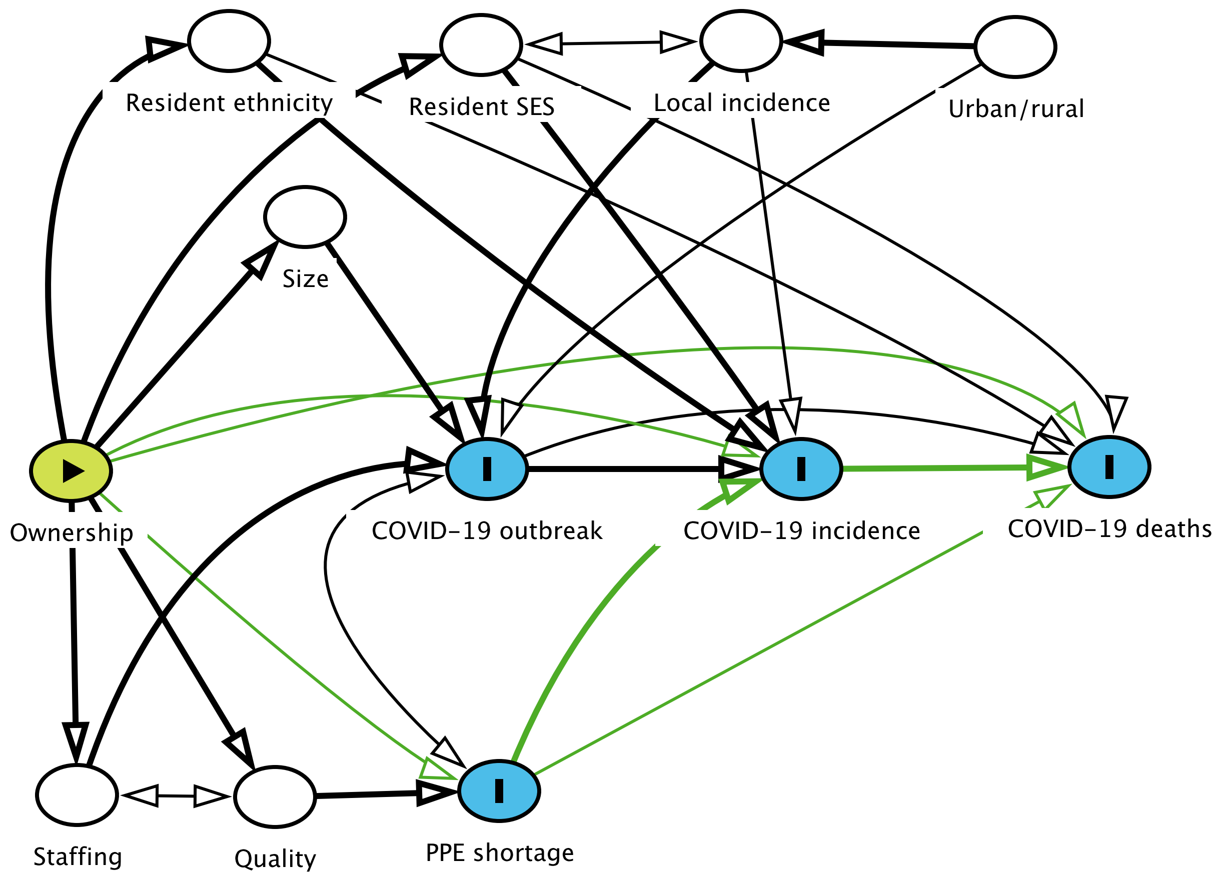
Resident ethnicity
Resident SES
Local incidence
Urban/rural
Size
Ownership
COVID−19 outbreak
COVID−19 incidence
COVID−19 deaths
Staffing
Quality
PPE shortage
